# MRI-derived body composition in bipolar and psychotic disorders

**DOI:** 10.64898/2025.12.10.25341959

**Authors:** Daniel E. Askeland-Gjerde, Lars T. Westlye, Dag Alnæs, Patrik Andersson, Dani Beck, Ingrid T. Johansen, Jennifer Linge, Geneviève Richard, Henning S. Rise, Paul M. Thompson, Sigrun Halvorsen, Ole A. Andreassen, Tiril P. Gurholt

**Affiliations:** Section for Precision Psychiatry, Division of Mental Health and Addiction, Oslo University Hospital, Oslo, Norway; Institute of Clinical Medicine, University of Oslo, Oslo, Norway; Department of Psychology, University of Oslo, Oslo, Norway; KG Jebsen Centre for Neurodevelopmental Disorders, University of Oslo, Oslo, Norway; AMRA Medical AB, Linköping, Sweden; PROMENTA Research Center, Department of Psychology, University of Oslo, Oslo, Norway; Department of Health, Medicine and Caring Sciences, Linköping University, Linköping, Sweden; Institute of Basic Medical Sciences, Division of Anatomy, University of Oslo, Oslo, Norway; Department of Neurology, Oslo University Hospital, Oslo, Norway; Imaging Genetics Center, Mark & Mary Stevens Neuroimaging & Informatics Institute, Keck School of Medicine, University of Southern California, Marina del Rey, CA, USA; Department of Cardiology, Oslo University Hospital Ullevål, Oslo, Norway

## Abstract

**Objective:** Bipolar (BD) and psychotic disorders (PD) are severe mental disorders (SMDs) associated with higher cardiometabolic disease burden. Prior studies of body composition in SMDs are inconclusive. Magnetic resonance imaging (MRI) offers detailed insights into body composition and may improve our understanding of cardiometabolic disease risk in patients with SMDs.

**Methods:** We derived body composition measures from MRI of patients with BD (n=36, 63.9% females, mean age=36.6±10.9 years), PD (n=27, 33.3% females, mean age=31.7±8.9 years), and controls (n=251, 78.8% females, mean age=40.7±12.1 years). Using linear models we tested for case-control differences in body mass index (BMI) and body composition in all patients and split by diagnostic group (BD and PD), accounting for age, sex, and scanner, and subsequently adding BMI. Next, we tested for age- and sex-adjusted associations between body composition and antipsychotic dose in patients.

**Results:** Analyses showed significantly higher BMI (BD: Cohen’s d=0.70; PD: d=0.65), abdominal subcutaneous fat (BD: d=0.78; PD: d=0.71), visceral fat (BD: d=0.76; PD=0.68), liver fat (BD: d=0.53; PD: n.s.), and thigh muscle fat infiltration (BD: d=0.80; PD: d=0.87) in cases compared to controls. When accounting for BMI, muscle fat infiltration remained significantly higher (BD: d=0.54; PD: d=0.68) and thigh muscle volume was lower (BD: d=-0.65; PD: d=-0.63). There were no significant associations of antipsychotic dose with BMI or body composition.

**Conclusions:** The observed case-control differences suggest that individuals with BD and PD, at the group level, exhibit higher regional fat accumulation and lower muscle volume, possibly contributing to the heightened risk for cardiometabolic comorbidity in SMDs.

## Introduction

Patients with severe mental disorders (SMDs) such as bipolar disorder (BD) and psychotic disorders (PD) have a higher risk of premature death (1), largely attributed to cardiometabolic disease (2). Body composition measures derived from imaging methods, such as magnetic resonance imaging (MRI), have yielded insight into cardiometabolic abnormalities and have been suggested as sensitive markers of cardiometabolic risk beyond conventional measures, such as body mass index (BMI), in community-dwelling participants and cardiometabolic patients (3–5). Prior reports of higher glucose, lipids, and inflammatory markers in patients with SMDs compared to controls suggest an elevated risk of adverse body composition (6–8), but previous studies on imaging-derived body composition measures are inconclusive (9–11). Therefore, little is known about body composition differences in patients with BD and PD relative to controls and how this relates to psychopharmacological treatment.

The regulation of body fat distribution is complex (12). In healthy individuals, lipids are primarily stored subcutaneously (12). However, in chronic overnutrition and in disease states, subcutaneous storage capacity can be exceeded, putatively due to adipocyte hypertrophy and insulin resistance (12). Exceeded subcutaneous storage capacity may lead to visceral and ectopic fat expansion, including liver fat and muscle fat infiltration (12). The classical anthropometric measures do not provide information on body fat and muscle composition, leading to a loss of clinically relevant information as the distribution of body fat compartments and muscle mass are differentially associated with cardiometabolic disease (13). Body MRI allows for direct and detailed measures of body composition (14), which have been linked to brain health and brain aging in community-dwelling adults (15–18), and may serve as sensitive measures for screening and monitoring of cardiometabolic burden in patients with SMDs (19).

Current literature indicates that patients with BD and PD often have elevated BMI and higher obesity prevalence compared to the general population (6), contributing to a higher risk of cardiometabolic disease (6). These observations have been linked to both the effects of antipsychotic medication (6), lifestyle factors (20), and common genetic factors (21,22), suggesting a multifactorial basis for weight gain and metabolic disease in these conditions. Previous studies of body composition in BD and PD have, however, shown mixed results. The only study on direct body composition measures in BD reported no differences in computed tomography-derived abdominal subcutaneous fat, visceral fat, or muscle fat infiltration in women with BD I compared to matched controls (9). A meta-analysis of studies of body composition that used dual energy x-ray absorptiometry, computed tomography, or MRI reported higher subcutaneous but not visceral fat in patients with schizophrenia relative to controls (11). Conversely, an MRI study showed no differences in total or visceral fat in schizophrenia patients compared to matched controls (10). To summarize, there are few studies of direct body composition measures in patients with BD and PD and they are mostly inconclusive. Prior studies suggest that antipsychotic treatment is associated with increased subcutaneous and visceral fat in adults with schizophrenia and children with mental disorders (11,19). However, the relationship between antipsychotic treatment and other body composition measures remains untested.

To address these knowledge gaps, we leveraged body MRI data gathered from a single site to test for case-control differences in body composition measures in patients with BD and PD, and to investigate their relationships with antipsychotic dose verified by serum concentration measures. Based on the literature reviewed above, we hypothesized that patients with BD and PD would have higher levels of BMI, subcutaneous, visceral, and ectopic fat and lower muscle volume than controls, and that antipsychotic treatment would be associated with higher BMI and body fat. We first tested for case-control differences in body composition in BD and PD, both combined and separately, using models first without and subsequently with BMI as a covariate. Second, we tested for associations between antipsychotic dose and body composition among patients.

## Methods and materials

### Participants

Participants aged 18 to 65 years with body MRI data obtained from the Regional Core Facility in Translational MRI Neuroimaging at Oslo University Hospital, Norway, were eligible for inclusion. In total, we included 314 participants. This included SMD patients consisting of BD (n=36) and PD (n=27) from the Thematically Organized Psychosis (TOP) study, and 251 controls from the TOP, StrokeMRI, and BRAINMINT studies. Patients were recruited from outpatient and inpatient psychiatric clinics in the Oslo region. Controls were recruited from the same area and invited by random draw from national records or newspaper and social media advertising. In this study, control eligibility criteria included no history of mental or neurological disorders. A subset of controls was assessed with Primary Care Evaluation of Mental Disorder (PRIME-MD) (24) and reported the medical history of first-degree relatives. When applicable, controls who had first-degree relatives with SMDs were excluded (n=45). We excluded patients and controls with a history of cardiovascular disease (n=2) or diabetes (n=7). Patients with a history of moderate to severe head injury, neurological or organic brain disorders, or intellectual developmental disorder were also excluded (although there were none in the initial sample; n=0). Additionally, controls who lacked data on body composition measures, height, or weight were excluded (n=33). All participants provided written informed consent. The current study was approved by the Regional Committee for Medical Research Ethics South East Norway (REC: 2009/2485, 2014/694, 2019/345) and conducted in accordance with the Declaration of Helsinki.

### Clinical information

Patients were assessed by physicians or clinical psychologists with the Structured Clinical Interview (SCID-I) for the Diagnostic and Statistical Manual of Mental Disorders (DSM-IV) and the Positive and Negative Syndrome Scale (PANSS) (23,24). We grouped patients into BD, including BD I (n=15) and BD II (n=21), and PD, including schizophrenia (n=9), schizophreniform disorder (n=3), schizoaffective disorder (n=7), delusional disorder (n=3), and PD not otherwise specified (n=5).

Height and weight were measured for all participants, and fasting blood samples were drawn from patients and a subset of controls. We calculated BMI (kg/m^2^) and classified participants into underweight (BMI<18.5), normal weight (18.5≤BMI<25), overweight (25≤BMI<30), and obesity (BMI≤30). Participants self-reported their assigned sex at birth. Blood samples were analyzed at the Department of Medical Biochemistry, Oslo University Hospital, and analyses included glycated hemoglobin, glucose, C-reactive protein, high-density lipoprotein cholesterol, low-density lipoprotein cholesterol, total cholesterol, and triglycerides.

We obtained psychopharmacological treatment information from interviews and medical records, and grouped medication into antipsychotics, lithium, antiepileptics, and antidepressants. Thirty-six (58.1%) patients reported current use of antipsychotics, almost exclusively second-generation agents, most frequently aripiprazole (n=12), olanzapine (n=10), and quetiapine (n=9). Five (8.1%) patients used multiple antipsychotics, and 28 (45.2%) used multiple psychotropic drugs. Seven (11.3%) patients reported using lithium, 19 (30.6%) antiepileptics, and 17 (27.4%) antidepressants. We standardized doses of antipsychotics using Defined Daily Dose (https://www.whocc.no/atc_ddd_index/), and in the case of antipsychotic polypharmacy, summed the Defined Daily Doses. The Department of Clinical Pharmacology, St. Olav University Hospital, Trondheim, Norway analyzed psychopharmacological serum concentrations. Eleven patients who reported using antipsychotics lacked serum concentrations, either because they refused to give blood samples, due to technical issues with the sample analysis, or due to very low dose adjunctive treatment. Serum concentrations below the dose-adjusted reference range (n=2) were defined as missing.

### MRI Acquisition and Processing

All participants underwent 3T body MRI at the Regional Core Facility in Translational MRI Neuroimaging at Oslo University Hospital, Norway between April 2018 and September 2023. The 3T GE Discovery MR750 was replaced with a 3T GE Signa Premier in December 2020. Controls were scanned before (n=152) and after (n=99), while patients were only scanned after the scanner upgrade. Identical sequences and parameters were used for both scanners. LAVA Flex was used to acquire water and fat separated data in the area between the neck and knees with minimum full echo time, 10° flip angle, 50 × 50 mm field of view, 5 mm slice thickness, and total scan time of approximately 6:00 minutes. IDEAL IQ sequence was used for liver fat assessment with minimum echo time, 3° flip angle, 45 × 45 mm field of view, 8 mm slice thickness, and 0:22 minutes scan time.

AMRA Researcher (AMRA Medical, Linköping, Sweden) was applied to derive body composition measures from the body MRI data, including measures of abdominal subcutaneous fat, visceral fat, liver fat, muscle fat infiltration, and muscle volume. The analysis is described in detail in previous publications (5,25). In brief, AMRA calibrated the images, fused the image stacks, segmented the resultant images, and quantified fat and muscle volumes (5). Abdominal subcutaneous fat and visceral fat are defined as fat in the subcutis from the top of the femoral head to the top of the 9^th^ thoracic vertebra and fat inside the abdominal cavity, respectively (5). Liver fat is computed by averaging the proton density fat fraction from 3-9 regions of interest in the liver, avoiding inhomogeneities, major vessels, and bile ducts (5). Muscle fat infiltration and muscle volume are defined as the mean fat fraction and muscle volume of the viable (fat fraction <50%) thigh muscle tissue, respectively (5).

### Statistical analyses

All analyses were conducted in R version 4.3.2 (26) with R-packages *tidyverse*, *RColorBrewer*, *MASS*, *MBESS*, *emmeans*, *ggplot2*, and *patchwork*. Continuous variables were assessed with quantile-quantile plots, histograms, and correlation plots (**Figure 1**, **Figures S1-S2**) to evaluate distributional properties and detect potential outliers or collinearity. We averaged right, left, anterior, and posterior muscle fat infiltration and muscle volume measures. Differences in continuous and categorical variables between the different groups (controls, all patients, bipolar disorder, psychotic disorder) were tested using two-sided t-tests and chi-squared tests, respectively (**Tables 1-2, Tables S1-S3**).

**Figure 1:**
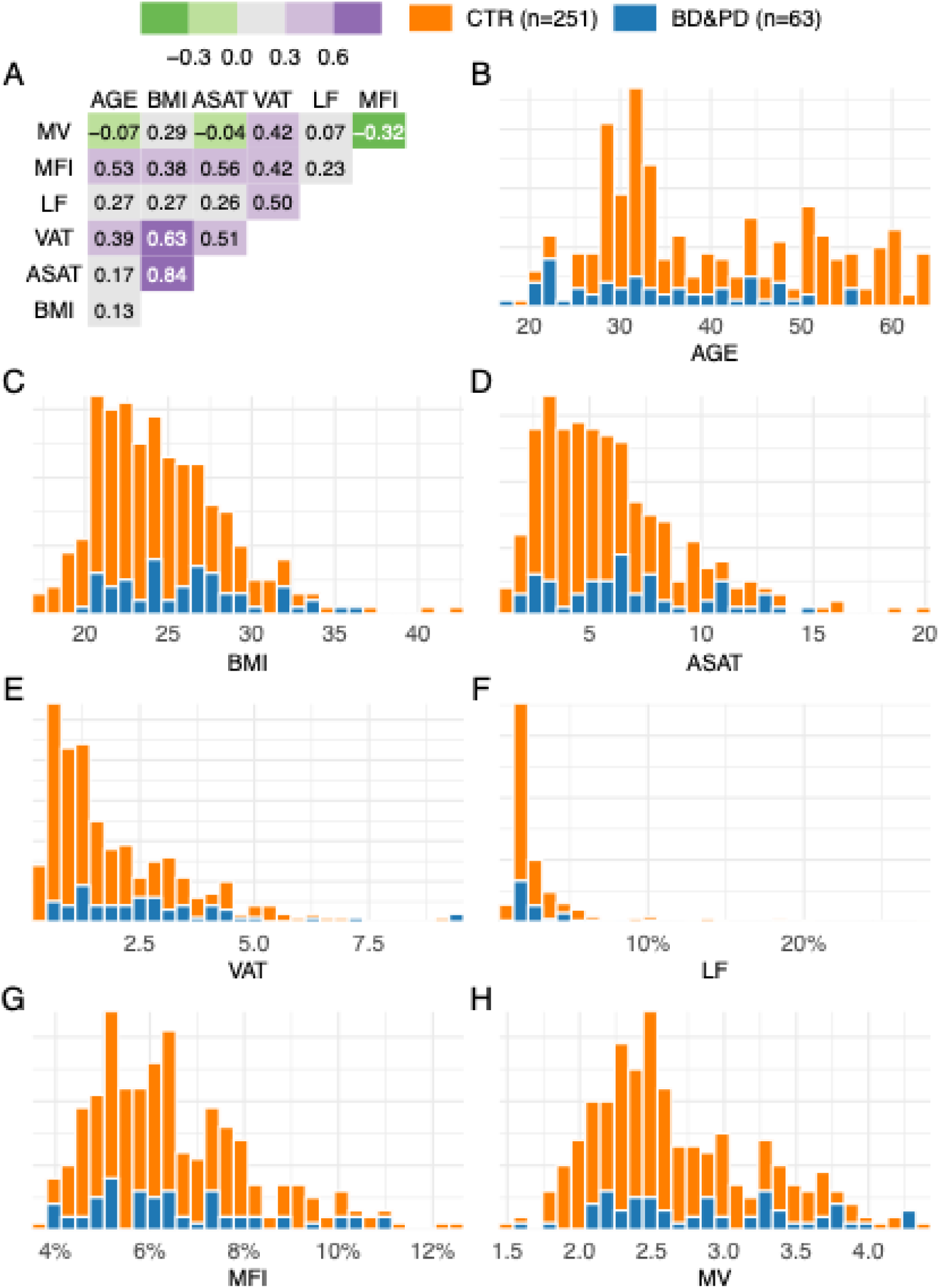
Correlations and distributions of age, body mass index, and body composition. **A.** The correlations of age, body mass index (BMI), abdominal subcutaneous adipose tissue (ASAT), visceral adipose tissue (VAT), liver fat (LF), muscle fat infiltration (MFI), and muscle volume (MV). **B-H.** The distributions of age, BMI, and ASAT, VAT, LF, MFI, and MV among controls (CTR) and patients (BD&PD). **Figure 1 alt text:** Heatmap showing stronger positive correlations of body mass index with abdominal subcutaneous fat and visceral fat than with liver fat, muscle fat infiltration, and muscle volume. Additionally, histograms depicting the distribution of age, body mass index, and the body composition measures for study participants split into the patient and control group, generally showing non-normal distributions of body composition measures and higher body mass index, abdominal subcutaneous fat, visceral fat, and muscle volume in the patient group.

**Table 1:**
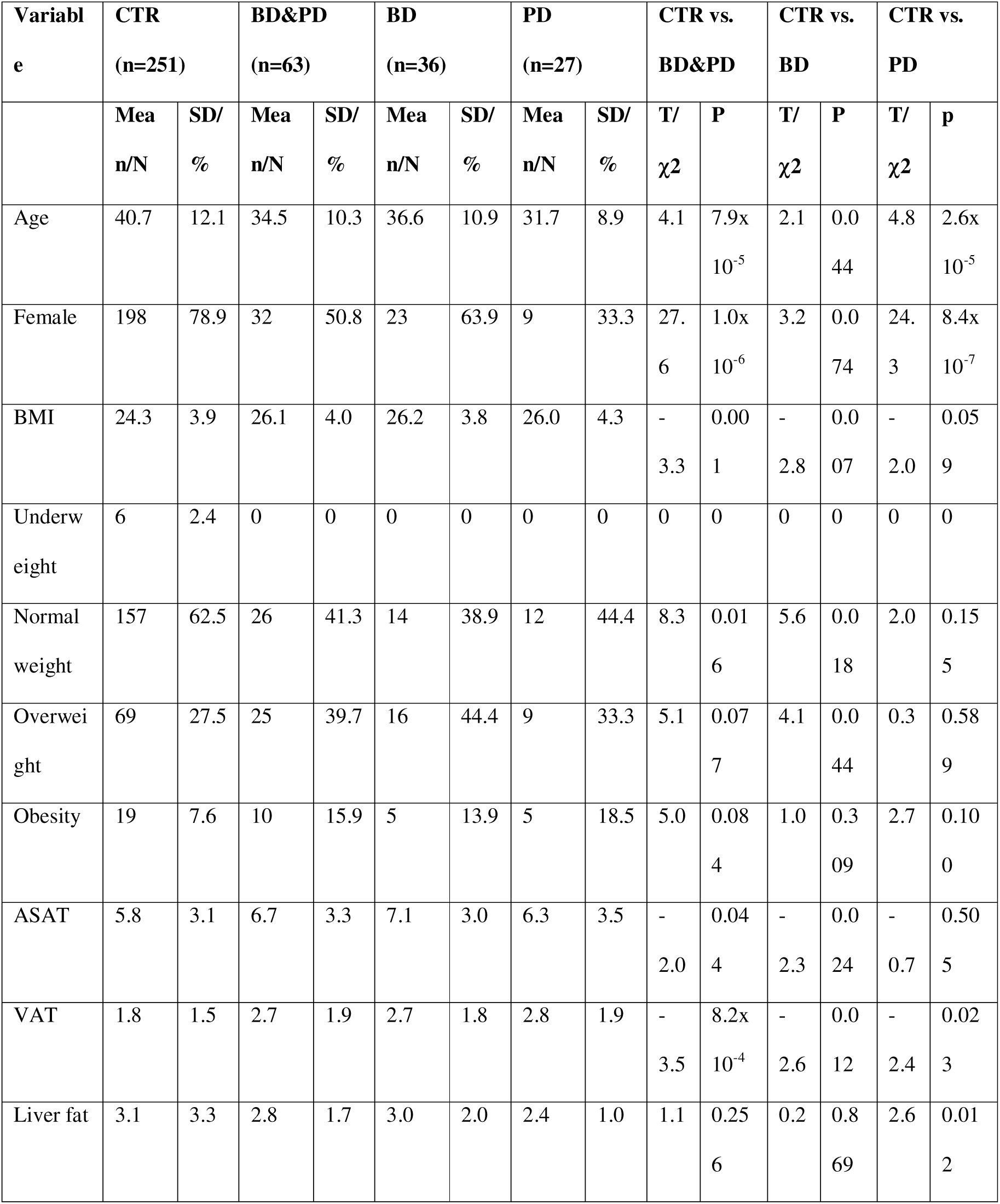
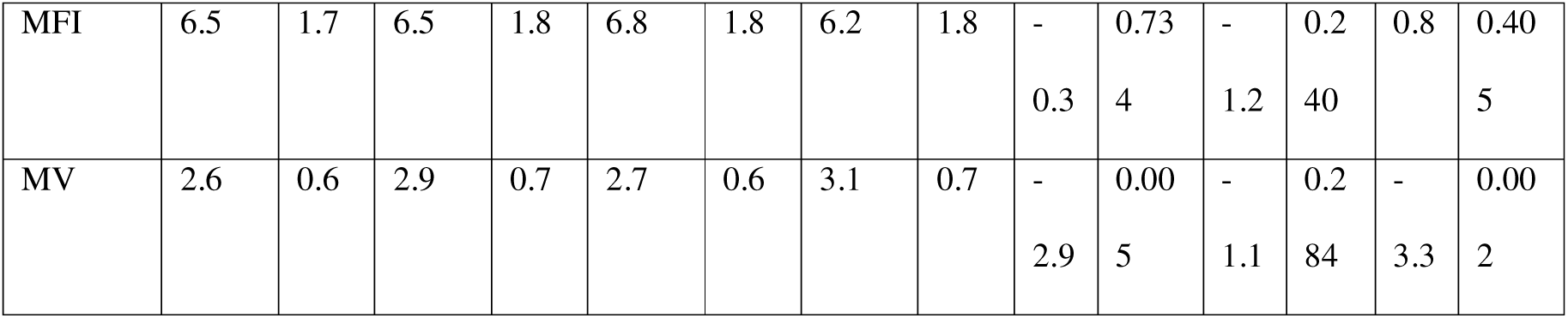
Sample characteristics: Sample characteristics and tests for differences between controls (CTR) and all patients (BD&PD), between controls and patients with bipolar disorder (BD), and between controls and patients with psychotic disorder (PD). We used t-tests for continuous and chi-squared tests for categorical measures. Statistically significant p-values are in bold. *Abbreviations*: BMI=body mass index, ASAT=abdominal subcutaneous adipose tissue, VAT=visceral adipose tissue, MFI=muscle fat infiltration, MV=muscle volume. *Missing data points:* BD: BMI (n=1), MFI (n=1), MV (n=1); PD: BMI (n=1), LF (n=3).

**Table 2:**
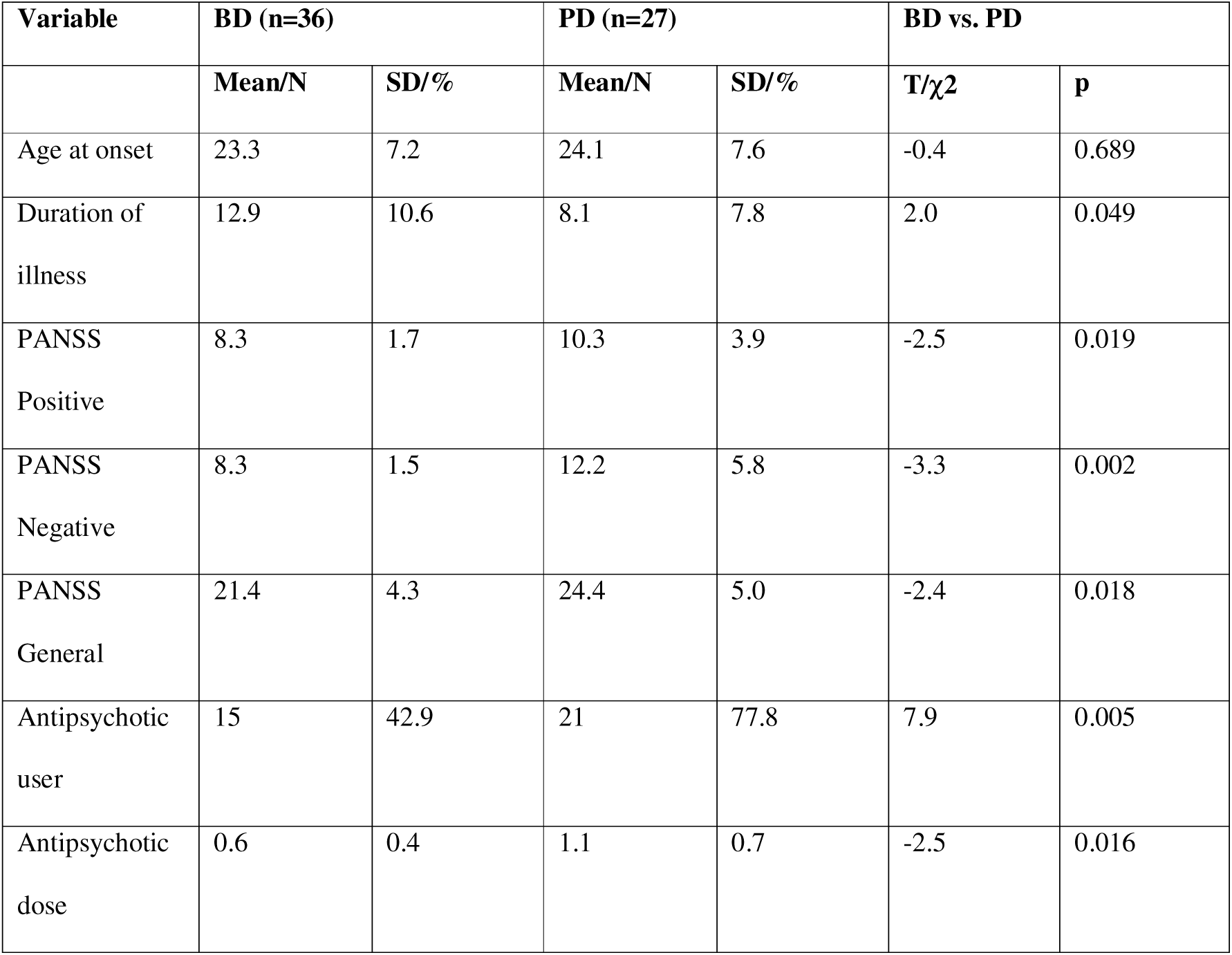
Patient sample characteristics: Patient sample characteristics and tests for differences between patients with bipolar disorder (BD) and patients with psychotic disorder (PD). We used t-tests for continuous and chi-squared tests for categorical measures. Statistically significant p-values are in bold. *Abbreviations*: PANSS=positive and negative syndrome scale. *Missing data points*: BD: age at onset (n=3), duration of illness (n=3), PANSS (n=3); PD: age at onset (n=1), duration of illness (n=1), PANSS (n=3), antipsychotic user (n=1).

| Variable | BD (n=36) |  | PD (n=27) |  | BD vs. PD |  |
| --- | --- | --- | --- | --- | --- | --- |
| | Mean/N | SD/% | Mean/N | SD/% | T/$\chi^2$ | p |
| Age at onset | 23.3 | 7.2 | 24.1 | 7.6 | -0.4 | 0.689 |
| Duration of illness | 12.9 | 10.6 | 8.1 | 7.8 | 2.0 | 0.049 |
| PANSS Positive | 8.3 | 1.7 | 10.3 | 3.9 | -2.5 | 0.019 |
| PANSS Negative | 8.3 | 1.5 | 12.2 | 5.8 | -3.3 | 0.002 |
| PANSS General | 21.4 | 4.3 | 24.4 | 5.0 | -2.4 | 0.018 |
| Antipsychotic user | 15 | 42.9 | 21 | 77.8 | 7.9 | 0.005 |
| Antipsychotic dose | 0.6 | 0.4 | 1.1 | 0.7 | -2.5 | 0.016 |

Age, sex, and MRI scanner differences can influence body composition measures (25,27). It should however be noted that the methodology applied in AMRA Researcher has been shown to be independent of scanner differences (25). We opted to adjust all multiple linear regression models for age and sex and, when relevant, scanner version. We tested for case-control differences in body composition measures in 1) all patients, and 2) patients split into BD and PD, compared to controls. These analyses were performed with and without BMI as a covariate. Next, we investigated associations between antipsychotic dose and body composition measures in 1) all patients, and 2) all patients except those whose antipsychotic use could not be verified by serum concentration measures. We log-transformed BMI, abdominal subcutaneous fat, visceral fat, liver fat, and muscle fat infiltration due to non-normal residuals in the regression models. As the residuals were still non-normal for log-transformed liver fat, we used Box-Cox transformation (5).

We calculated Cohen’s d for categorical and partial correlation coefficient (r) for continuous predictors, and computed 95% confidence intervals for each measure. We adjusted for multiple comparisons with a false discovery rate (FDR; q=0.05) using the Benjamini-Hochberg method (28) and reported adjusted p-values.

## Results

### Sample characteristics

Patients were, on average, younger (mean age=34.5±10.3 years vs. mean age=40.7±12.1 years) and had a higher proportion of males compared to the control group (78.9% vs. 50.8%; **Table 1**). Unadjusted comparisons showed that patients on average had higher BMI (mean=26.1±4.0 kg/m^2^ vs. mean=24.3±3.9 kg/m^2^), abdominal subcutaneous fat (mean=6.7±3.3 L vs. mean=5.8±3.1 L), visceral fat (mean=2.7±1.9 L vs. mean=1.8±1.5 L), and muscle volume (mean=2.9±0.7 L vs. mean=2.6±0.6 L), and a lower likelihood of being normal weight (41.3% vs. 62.5%) than controls.

Comparisons between the diagnostic groups showed that patients with PD were more likely to use antipsychotics (77.8% vs. 42.9%) and had higher PANSS scores on average (Positive: mean=10.3±3.9 vs. 8.3±1.7; Negative: mean=12.2±5.8 vs. 8.3±1.5; General: mean=24.4±5.0 vs. 21.4±4.3), and used higher defined daily doses of antipsychotics (mean=1.1±0.7 vs. 0.6±0.4), but had a shorter duration of illness (mean=8.1±7.8 years vs. 12.9±10.6 years), than patients with BD (**Table 2**).

### Body composition measures in bipolar and psychotic disorders

Multiple linear regression analyses showed significantly higher BMI (d=0.68, p=0.001), abdominal subcutaneous fat (d=0.75, p=6.3×10^-4^), visceral fat (d=0.73, p=7.9×10^-4^), liver fat (d=0.42, p=0.042), and muscle fat infiltration (d=0.83, p=2.5×10^-4^) in patients compared to controls (**Figure 2**, **Table 3**). When split by diagnostic group, both patients with BD and PD had higher BMI (BD: d=0.70, p=0.004; PD: d=0.65, p=0.020), abdominal subcutaneous fat (BD: 0.78, p=0.001; PD: d=0.71, p=0.010), visceral fat (BD: d=0.76, p=0.002; PD: d=0.68, p=0.015), and muscle fat infiltration (BD: d=0.80, p=0.001; PD: d=0.87, p=0.002) than controls. Additionally, BD patients had higher liver fat relative to controls (d=0.53, p=0.025).

**Figure 2:**
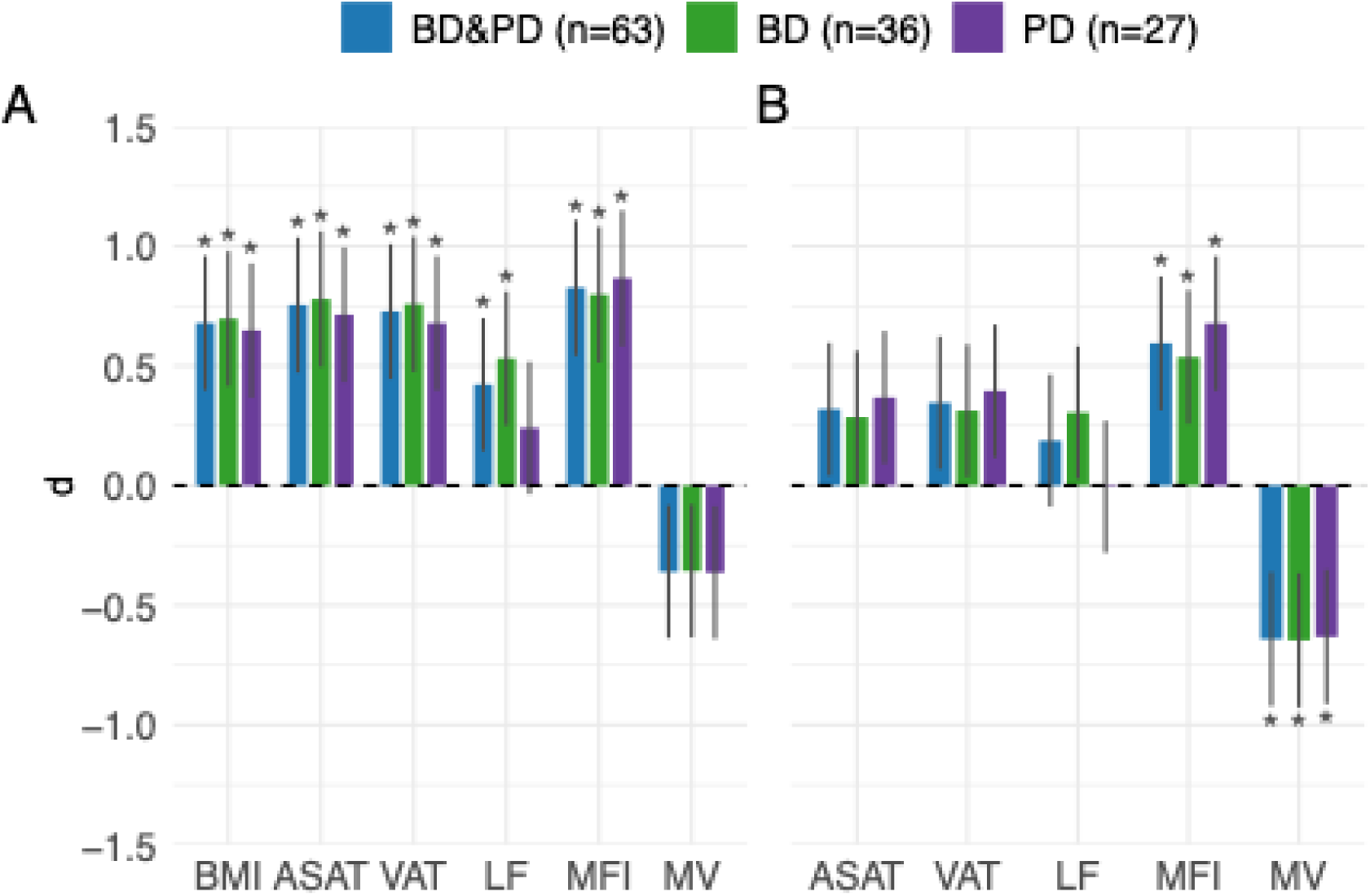
Case-control differences in body composition. **A.** The difference in body mass index (BMI), abdominal subcutaneous adipose tissue (ASAT), visceral adipose tissue (VAT), liver fat (LF), muscle fat infiltration (MFI), and muscle volume (MV) in all patients (BD&PD), patients with bipolar disorder (BD), and patients with psychotic disorder (PD) relative to controls. Adjusted for age, sex, and scanner. The lines correspond to 95% confidence intervals and asterisks (*) indicate statistical significance after adjustment for multiple comparisons. **B.** The difference in ASAT, VAT, LF, MFI, and MV in BD&PD, BD, and PD relative to controls additionally adjusted for BMI. *Abbreviations*: d=Cohen’s d. **Figure 2 alt text:** Two sets of bar charts depicting significantly higher body mass index and body fats in patients with bipolar and psychotic disorders, and after accounting for body mass index, higher muscle fat infiltration and lower muscle volume.

**Table 3:** Case-control differences in body composition. The difference in body mass index (BMI), abdominal subcutaneous adipose tissue (ASAT), visceral adipose tissue (VAT), liver fat (LF), muscle fat infiltration (MFI), and muscle volume (MV) in all patients (BD&PD), bipolar disorder (BD), and psychotic disorder (PD) compared to controls. The partially adjusted model included age, sex, and scanner as covariates. The fully adjusted model additionally included BMI as a covariate. Statistically significant adjusted p-values are in bold. *Abbreviations*: β=regression coefficient, SE=standard error, T=t-statistic, d=Cohen’s d, CI=confidence interval.

| Partially adjusted model |  |  |  |  |  |  |
| --- | --- | --- | --- | --- | --- | --- |
| Outcome | $\beta$ | SE | T | d | 95% CI | p |
| BD&PD (n=63) |  |  |  |  |  |  |
| BMI | 0.10 | 0.03 | 3.36 | 0.70 | 0.40, 0.96 | 0.001 |
| ASAT | 0.32 | 0.08 | 3.81 | 0.78 | 0.47, 1.04 | $6.3 \times 10^{-4}$ |
| VAT | 0.27 | 0.07 | 3.70 | 0.76 | 0.45, 1.01 | $7.9 \times 10^{-4}$ |
| LF | 21.91 | 8.45 | 2.59 | 0.53 | 0.14, 0.70 | 0.042 |
| MFI | 0.01 | 0.00 | 3.85 | 0.80 | 0.54, 1.11 | $2.5 \times 10^{-4}$ |
| MV | -0.13 | 0.08 | -1.72 | -0.36 | -0.64, -0.08 | 0.096 |
| BD (n=36) |  |  |  |  |  |  |
| BMI | 0.10 | 0.03 | 3.36 | 0.70 | 0.42, 0.98 | 0.004 |
| ASAT | 0.32 | 0.08 | 3.81 | 0.78 | 0.50, 1.06 | 0.001 |
| VAT | 0.27 | 0.07 | 3.70 | 0.76 | 0.48, 1.04 | 0.002 |
| LF | 21.91 | 8.45 | 2.59 | 0.53 | 0.25, 0.81 | 0.025 |
| MFI | 0.01 | 0.00 | 3.85 | 0.80 | 0.51, 1.08 | 0.001 |
| MV | -0.13 | 0.08 | -1.72 | -0.36 | -0.63, -0.08 | 0.138 |
| PD (n=27) |  |  |  |  |  |  |
| BMI | 0.09 | 0.03 | 2.70 | 0.65 | 0.37, 0.93 | 0.020 |
| ASAT | 0.29 | 0.10 | 3.01 | 0.71 | 0.43, 1.00 | 0.010 |
| VAT | 0.24 | 0.08 | 2.86 | 0.68 | 0.40, 0.96 | 0.015 |
| LF | 9.94 | 10.12 | 0.98 | 0.24 | -0.04, 0.52 | 0.350 |
| MFI | 0.01 | 0.00 | 3.65 | 0.87 | 0.58, 1.15 | 0.002 |
| MV | -0.13 | 0.09 | -1.53 | -0.36 | -0.64, -0.09 | 0.183 |
| Fully adjusted model |  |  |  |  |  |  |
| Outcome | $\beta$ | SE | T | d | 95% CI | p |
| BD&PD (n=63) |  |  |  |  |  |  |
| ASAT | 0.07 | 0.04 | 1.74 | 0.32 | 0.04, 0.60 | 0.138 |
| VAT | 0.08 | 0.04 | 1.89 | 0.35 | 0.07, 0.62 | 0.118 |
| LF | 7.27 | 7.16 | 1.02 | 0.19 | -0.09, 0.46 | 0.341 |
| MFI | 0.01 | 0.00 | 3.22 | 0.59 | 0.31, 0.87 | 0.006 |
| MV | -0.22 | 0.06 | -3.48 | -0.64 | -0.92, -0.36 | 0.003 |
| **BD (n=36)** |  |  |  |  |  |  |
| ASAT | 0.06 | 0.05 | 1.35 | 0.29 | 0.01, 0.56 | 0.216 |
| VAT | 0.08 | 0.05 | 1.48 | 0.31 | 0.04, 0.59 | 0.185 |
| LF | 11.79 | 8.18 | 1.44 | 0.31 | 0.03, 0.58 | 0.193 |
| MFI | 0.01 | 0.00 | 2.51 | 0.54 | 0.26, 0.82 | 0.030 |
| MV | -0.22 | 0.07 | -3.03 | -0.65 | -0.93, -0.37 | 0.010 |
| **PD (n=27)** |  |  |  |  |  |  |
| ASAT | 0.08 | 0.05 | 1.52 | 0.37 | 0.09, 0.65 | 0.183 |
| VAT | 0.10 | 0.06 | 1.63 | 0.40 | 0.12, 0.67 | 0.157 |
| LF | -0.21 | 9.70 | -0.02 | -0.01 | -0.28, 0.27 | 0.986 |
| MFI | 0.01 | 0.00 | 2.79 | 0.68 | 0.40, 0.96 | 0.017 |
| MV | -0.22 | 0.08 | -2.61 | -0.63 | -0.91, -0.35 | 0.025 |

Differences in abdominal subcutaneous fat, visceral fat, and liver fat between patients and controls were attenuated and no longer significant after including BMI in the model. However, muscle fat infiltration remained higher (d=0.59, p=0.006), and muscle volume was lower (d=-0.64, p=0.003) in patients compared to controls after BMI adjustment. Similar patterns were observed in analyses stratified by diagnostic groups. Both BD and PD patients had higher muscle fat infiltration (BD: d=0.54, p=0.030; PD: d=0.68, p=0.017) and lower muscle volume (BD: d=-0.93, p=0.010; PD: d=-0.91, p=0.025) than controls, while the differences in abdominal subcutaneous fat, visceral fat, and liver fat were attenuated and no longer statistically significant after adjusting for BMI.

### Antipsychotic dose and body composition measures

Multiple linear regression analyses revealed no significant associations between antipsychotic dose and body composition measures in the full patient sample nor in the patient sample without those whose antipsychotic use could not be verified by serum measures (**Table S2**).

## Discussion

We leveraged body MRI to demonstrate higher abdominal subcutaneous fat, visceral fat, and muscle fat infiltration in patients with BD and PD. The analyses showed that BMI was higher among patients, however, after adjusting for BMI, muscle fat infiltration remained higher and muscle volume was lower in patients compared to controls. These case-control differences suggest a higher risk of metabolically unhealthy body composition, characterized by increased visceral and ectopic fat and lower muscle mass, in patients with SMDs, which may play a role in the higher cardiometabolic disease burden in these conditions. Importantly, the higher risk of adverse muscle composition after accounting for BMI may go undetected as muscle composition is not included in conventional cardiometabolic assessment, potentially reflecting unmeasured cardiometabolic risk in patients with SMDs.

In line with our hypotheses, we observed similar patterns across diagnostic groups, namely, higher BMI, abdominal subcutaneous fat, visceral fat, and muscle fat infiltration with moderate to large effect sizes. The exception was liver fat, which was moderately higher in the combined patient group and in BD, but not significantly higher in PD, compared to controls. Liver fat accumulation can result from metabolic dysfunction but also from factors not accounted for in our analyses, such as excessive alcohol consumption and viral infections (29), which may have influenced our findings. Our findings demonstrate that patients with BD and PD have higher levels of visceral and ectopic fat, which have been associated with cardiometabolic disease beyond conventional cardiometabolic markers (13). However, case-control differences in subcutaneous, visceral, and liver fat were attenuated and no longer significant after adjustment for BMI. This is in accordance with most, but not all, studies on imaging-derived total, subcutaneous, and visceral fat in patients with SMDs (9–11). Interestingly, in models with BMI as a covariate, muscle fat infiltration remained higher and muscle volume was lower in all patient groups relative to controls. To our knowledge, only one prior study has investigated imaging-derived muscle fat infiltration in patients with SMD (9), and no prior studies have investigated imaging-derived muscle volume. Differences in sample size and characteristics, as well as variation in imaging and statistical methods, may have contributed to the null findings in the former study on muscle fat infiltration, contrasted with the significantly higher muscle fat infiltration observed among patients in this study. The observed moderately lower muscle quality and quantity may in part be related to lower rates of physical activity among patients with BD and PD (20). Worryingly, poor muscular health often goes undetected, especially in individuals with concurrent overweight or obesity (30), and is associated with cardiometabolic morbidity and mortality (30,31). Our findings suggest that impaired muscle quality and quantity, not captured by classical anthropometric measures, may be linked to the elevated cardiometabolic burden in patients with SMDs.

Contrary to our hypothesis, based on prior findings of increased insulin resistance and subcutaneous and visceral fat following antipsychotic treatment (11,19), our analysis did not reveal significant associations of antipsychotic dose and BMI or body composition measures. Several factors may explain this. We investigated cross-sectional associations, but current antipsychotic non-users were not necessarily antipsychotic-naïve. Furthermore, clinicians may prescribe medications with lower metabolic risk to patients with BD and elevated cardiometabolic risk, and patients who experience metabolic side effects may switch to alternative treatments. Implementing sensitive body composition measures in randomized controlled trials of adults treated with antipsychotics may improve our understanding of the metabolic consequences of these treatments (19).

The metabolic abnormalities underlying the current findings of higher body fat levels in BD and PD are likely multidimensional and complex (12). The lipid storage capacity of subcutaneous adipose tissue varies among individuals and is influenced by adipocyte differentiation from progenitor cells (32). Genetic studies on both schizophrenia and insulin resistance have revealed associations with N-acetyltransferase 2 (NAT2), a gene involved in adipocyte differentiation (12,33). In individuals who gain weight but lack sufficient differentiation of adipocyte progenitor cells, lipids are stored in fewer adipocytes (32). This leads to adipocyte hypertrophy, which is suggested to have detrimental effects on cardiometabolic health (13). Adipocyte size studies are currently lacking in BD and PD. However, our findings lead us to speculate that patients with SMDs are at an increased risk of adipocyte hypertrophy and, consequently visceral and ectopic fat expansion.

Insulin resistance is closely linked to unhealthy fat deposition (12). The first sign of systemic insulin resistance is reduced glycogen synthesis in skeletal muscle after food intake, related to alterations in glycogen synthase kinase-3 (GSK3) (12). GSK3 is a potential lithium target and is related to loss-of-function variants implicated in BD and schizophrenia (34). Therefore, GSK3 may contribute to impaired insulin signaling observed in patients with BD and schizophrenia, as well as in first-degree relatives of patients with schizophrenia (35,36). Genetic and transcriptomic analyses on postmortem brain tissue from donors with schizophrenia provide converging evidence of mitochondrial dysregulation (37). Interestingly, genes differentially expressed between schizophrenia and controls were particularly enriched in the mitochondrial inner membrane (37), a key site of cellular metabolism. Genetic risk variants for BD and schizophrenia are enriched in enzymes related to the lipid diacylglycerol (38) and patients with BD and schizophrenia have higher serum levels of ceramides (38), which play an auxiliary role in diacylglycerol-induced insulin resistance (39). While findings from genomics, antipsychotic-naïve patients, and first-degree relatives suggest innate glucose abnormalities, antipsychotics also reduce insulin-stimulated muscle glucose uptake (19). Whether glucose dysregulation – innate or acquired – contributes to high-risk fat deposition in BD and PD remains to be determined.

Our results suggest that patients with BD and PD may be at higher risk of a metabolically unhealthy body composition. The observed higher burden of cardiometabolic disease in patients with SMDs is not fully explained by conventional cardiometabolic risk factors (6). Body composition measures have been associated with cardiometabolic disease beyond conventional cardiometabolic markers such as BMI, blood pressure, and serum measures in individuals without SMDs (3–5). Although similar associations might be expected in individuals with and without SMDs, these relationships remain untested in SMD populations. Still, we might speculate that metabolically unhealthy body composition may be an unassessed contributing factor to cardiometabolic risk in patients with SMDs. However, additional factors are likely involved in the development of cardiometabolic comorbidity in SMDs (6,20), which precludes definite conclusions.

Our study has both strengths and limitations. While we verified current antipsychotic treatment with serum concentration measures, the study was neither powered nor designed to differentiate antipsychotics based on their metabolic risk profiles. Patients were assessed, on average, in their mid-thirties and approximately 11 years after illness onset. While this allowed us to examine body composition years before the typical onset of cardiometabolic disease, our results may not generalize to patients with longer-lasting illness duration, in whom psychosocial burden, poor diet, and low physical activity may further impair cardiometabolic health. Nor do the findings necessarily generalize to antipsychotic-naïve patients as antipsychotics may influence body fat distribution (11,19). Race and ethnicity are considered relevant factors in cardiometabolic assessment (40). However, race and ethnicity were not consistently reported across cohorts and could therefore not be included as a covariate and thus our findings may not generalize. Finally, although we adjusted for sex in all analyses, unequal sex distribution across groups may have influenced our results.

In conclusion, our analyses revealed higher BMI, as well as elevated abdominal subcutaneous fat, visceral fat, liver fat, and muscle fat infiltration. When adjusting for BMI, higher muscle fat infiltration and lower muscle volume was observed in patients with BD and PD compared to controls. Together, these findings implicate adverse body composition in patients with SMDs and highlight the potential role of MRI-derived body composition measures as cardiometabolic risk markers in mental health care.

## Acknowledgements

Gottfried Gerkum and Halvor S. Devor, who both have lived experience expertise from severe mental illness, have provided input on the current work, with suggestions for improvements and future directions. We performed all data analyses on the Services for Sensitive Data, University of Oslo, Norway, with resources from UNINETT Sigma2 – the National Infrastructure for High-Performance Computing and Data Storage in Norway. TPG received funding from South-Eastern Norway Regional Health Authority (Grant No. 2022080), OAA from the European Union’s Horizon 2020 Research and Innovation Programme (CoMorMent project No. 802998) and Research Council of Norway (Grant No. 223273), and LTW from German Federal Ministry of Education and Research (COMMITMENT, Grant No. 01ZX1904A) and European Research Council StG (Grant No. 802998).

## Data availability

Due to privacy and ethical restrictions, the data utilized in the current study are not publicly available. However, upon reasonable request and after approval from the Regional Ethics Committee they can be made available from the corresponding author.

## Author contributions

**DEA-G:** Conceptualization, Methodology, Software, Formal analysis, Visualization, Data curation, Writing – Original Draft. **LTW:** Resources, Writing – Review & Editing, Supervision, Project administration, Funding acquisition. **DA:** Resources, Writing – Review & Editing, Project administration. **ASA:** Resources, Writing – Review & Editing. PA: Resources, Writing – Review & Editing. **DB:** Investigation, Data Curation, Writing – Review & Editing. **ITJ:** Investigation, Writing – Review & Editing. **JL:** Resources, Writing – Review & Editing. **GR:** Investigation, Data Curation, Writing – Review & Editing. **HSR:** Investigation, Data Curation, Writing – Review & Editing, Project administration. **PMT:** Writing – Review & Editing. **SH:** Writing – Review & Editing, Supervision. **OAA:** Conceptualization, Methodology, Resources, Writing – Review & Editing, Supervision, Project administration, Funding acquisition. **TPG:** Conceptualization, Methodology, Writing – Review & Editing, Supervision, Project administration, Funding acquisition.

## References

1. Plana-Ripoll O, Pedersen CB, Agerbo E, Holtz Y, Erlangsen A, Canudas-Romo V, et al. A comprehensive analysis of mortality-related health metrics associated with mental disorders: a nationwide, register-based cohort study. The Lancet. 2019 Nov 16;394(10211):1827–35.

2. Correll CU, Solmi M, Veronese N, Bortolato B, Rosson S, Santonastaso P, et al. Prevalence, incidence and mortality from cardiovascular disease in patients with pooled and specific severe mental illness: a large-scale meta-analysis of 3,211,768 patients and 113,383,368 controls. World Psychiatry. 2017;16(2):163–80.

3. Neeland IJ, Ross R, Després JP, Matsuzawa Y, Yamashita S, Shai I, et al. Visceral and ectopic fat, atherosclerosis, and cardiometabolic disease: a position statement. Lancet Diabetes Endocrinol. 2019 Sep 1;7(9):715–25.

4. Souza AC do AH, Troschel AS, Marquardt JP, Hadžić I, Foldyna B, Moura FA, et al. Skeletal muscle adiposity, coronary microvascular dysfunction, and adverse cardiovascular outcomes. Eur Heart J. 2025 Jan 20;ehae827.

5. Linge J, Borga M, West J, Tuthill T, Miller MR, Dumitriu A, et al. Body Composition Profiling in the UK Biobank Imaging Study. Obes Silver Spring Md. 2018 Nov;26(11):1785–95.

6. Polcwiartek C, O’Gallagher K, Friedman DJ, Correll CU, Solmi M, Jensen SE, et al. Severe mental illness: cardiovascular risk assessment and management. Eur Heart J. 2024 Mar 21;45(12):987–97.

7. Fernandes BS, Steiner J, Molendijk ML, Dodd S, Nardin P, Gonçalves CA, et al. C-reactive protein concentrations across the mood spectrum in bipolar disorder: a systematic review and meta-analysis. Lancet Psychiatry. 2016 Dec 1;3(12):1147–56.

8. Halstead S, Siskind D, Amft M, Wagner E, Yakimov V, Liu ZSJ, et al. Alteration patterns of peripheral concentrations of cytokines and associated inflammatory proteins in acute and chronic stages of schizophrenia: a systematic review and network meta-analysis. Lancet Psychiatry. 2023 Apr 1;10(4):260–71.

9. Fleet-Michaliszyn SB, Soreca I, Otto AD, Jakicic JM, Fagiolini A, Kupfer DJ, et al. A Prospective Observational Study of Obesity, Body Composition, and Insulin Resistance in 18 Women With Bipolar Disorder and 17 Matched Control Subjects. J Clin Psychiatry. 2008 Dec 31;69(12):15800.

10. Osimo EF, Brugger SP, Thomas EL, Howes OD. A cross-sectional MR study of body fat volumes and distribution in chronic schizophrenia. Schizophrenia. 2022 Mar 18;8(1):1–3.

11. Smith E, Singh R, Lee J, Colucci L, Graff-Guerrero A, Remington G, et al. Adiposity in schizophrenia: A systematic review and meta-analysis. Acta Psychiatr Scand. 2021;144(6):524–36.

12. Roden M, Shulman GI. The integrative biology of type 2 diabetes. Nature. 2019 Dec;576(7785):51–60.

13. Valenzuela PL, Carrera-Bastos P, Castillo-García A, Lieberman DE, Santos-Lozano A, Lucia A. Obesity and the risk of cardiometabolic diseases. Nat Rev Cardiol. 2023 Jul;20(7):475–94.

14. Neeland IJ, Yokoo T, Leinhard OD, Lavie CJ. 21st Century Advances in Multimodality Imaging of Obesity for Care of the Cardiovascular Patient. JACC Cardiovasc Imaging. 2021 Feb 1;14(2):482–94.

15. Beck D, de Lange AMG, Alnæs D, Maximov II, Pedersen ML, Leinhard OD, et al. Adipose tissue distribution from body MRI is associated with cross-sectional and longitudinal brain age in adults. NeuroImage Clin. 2022 Jan 27;33:102949.

16. Gurholt TP, Kaufmann T, Frei O, Alnæs D, Haukvik UK, van der Meer D, et al. Population-based body–brain mapping links brain morphology with anthropometrics and body composition. Transl Psychiatry. 2021 May 18;11(1):1–12.

17. Gurholt TP, Borda MG, Parker N, Forminykh V, Kjelkenes R, Linge J, et al. Linking sarcopenia, brain structure, and cognitive performance: a large-scale UK Biobank study. Brain Commun. 2024 Mar 7;fcae083.

18. Askeland-Gjerde DE, Westlye LT, Andersson P, Korbmacher M, Lange AM de, Meer D van der, et al. Mediation Analyses Link Cardiometabolic Factors and Liver Fat With White Matter Hyperintensities and Cognitive Performance: A UK Biobank Study. Biol Psychiatry Glob Open Sci [Internet]. 2025 Jul 1 [cited 2025 Jun 24];5(4). Available from: https://www.bpsgos.org/article/S2667-1743(25)00042-4/fulltext

19. Nicol GE, Yingling MD, Flavin KS, Schweiger JA, Patterson BW, Schechtman KB, et al. Metabolic Effects of Antipsychotics on Adiposity and Insulin Sensitivity in Youths: A Randomized Clinical Trial. JAMA Psychiatry. 2018 Aug 1;75(8):788–96.

20. Vancampfort D, Firth J, Schuch FB, Rosenbaum S, Mugisha J, Hallgren M, et al. Sedentary behavior and physical activity levels in people with schizophrenia, bipolar disorder and major depressive disorder: a global systematic review and meta-analysis. World Psychiatry. 2017;16(3):308–15.

21. Rødevand L, Rahman Z, Hindley GFL, Smeland OB, Frei O, Tekin TF, et al. Characterizing the Shared Genetic Underpinnings of Schizophrenia and Cardiovascular Disease Risk Factors. Am J Psychiatry. 2023 Nov;180(11):815–26.

22. Rødevand L, Bahrami S, Frei O, Chu Y, Shadrin A, O’Connell KS, et al. Extensive bidirectional genetic overlap between bipolar disorder and cardiovascular disease phenotypes. Transl Psychiatry. 2021 Jul 23;11(1):1–9.

23. First MB, Gibbon M. The Structured Clinical Interview for DSM-IV Axis I Disorders (SCID-I) and the Structured Clinical Interview for DSM-IV Axis II Disorders (SCID-II). In: Comprehensive handbook of psychological assessment, Vol 2: Personality assessment. Hoboken, NJ, US: John Wiley & Sons, Inc.; 2004. p. 134–43.

24. Kay SR, Fiszbein A, Opler LA. The Positive and Negative Syndrome Scale (PANSS) for Schizophrenia. Schizophr Bull. 1987 Jan 1;13(2):261–76.

25. Borga M, Ahlgren A, Romu T, Widholm P, Dahlqvist Leinhard O, West J. Reproducibility and repeatability of MRI-based body composition analysis. Magn Reson Med. 2020;84(6):3146–56.

26. R Core Team. R: A Language and Environment for Statistical Computing [Internet]. Vienna, Austria: R Foundation for Statistical Computing; 2024. Available from: https://www.R-project.org/

27. Goossens GH, Jocken JWE, Blaak EE. Sexual dimorphism in cardiometabolic health: the role of adipose tissue, muscle and liver. Nat Rev Endocrinol. 2021 Jan;17(1):47–66.

28. Benjamini Y, Hochberg Y. Controlling the False Discovery Rate: A Practical and Powerful Approach to Multiple Testing. J R Stat Soc Ser B Methodol. 1995;57(1):289–300.

29. Huang DQ, Wong VWS, Rinella ME, Boursier J, Lazarus JV, Yki-Järvinen H, et al. Metabolic dysfunction-associated steatotic liver disease in adults. Nat Rev Dis Primer. 2025 Mar 6;11(1):1–25.

30. Damluji AA, Alfaraidhy M, AlHajri N, Rohant NN, Kumar M, Al Malouf C, et al. Sarcopenia and Cardiovascular Diseases. Circulation. 2023 May 16;147(20):1534–53.

31. Linge J, Petersson M, Forsgren MF, Sanyal AJ, Dahlqvist Leinhard O. Adverse muscle composition predicts all-cause mortality in the UK Biobank imaging study. J Cachexia Sarcopenia Muscle. 2021;12(6):1513–26.

32. Lecoutre S, Rebière C, Maqdasy S, Lambert M, Dussaud S, Abatan JB, et al. Enhancing adipose tissue plasticity: progenitor cell roles in metabolic health. Nat Rev Endocrinol. 2025 Jan 6;1–17.

33. Trubetskoy V, Pardiñas AF, Qi T, Panagiotaropoulou G, Awasthi S, Bigdeli TB, et al. Mapping genomic loci implicates genes and synaptic biology in schizophrenia. Nature. 2022 Apr;604(7906):502–8.

34. Thorgeirsson TE, Tragante V, Sveinbjornsson G, Jonsdottir GA, Walters GB, Ivarsdottir EV, et al. Rare loss-of-function variants in HECTD2 and AKAP11 confer risk of bipolar disorder. Nat Genet. 2025 Apr;57(4):851–5.

35. Calkin CV, Ruzickova M, Uher R, Hajek T, Slaney CM, Garnham JS, et al. Insulin resistance and outcome in bipolar disorder. Br J Psychiatry. 2015 Jan;206(1):52–7.

36. Chouinard VA, Henderson DC, Dalla Man C, Valeri L, Gray BE, Ryan KP, et al. Impaired insulin signaling in unaffected siblings and patients with first-episode psychosis. Mol Psychiatry. 2019 Oct;24(10):1513–22.

37. Bast L, Yao S, Martínez-López JA, Memic F, French H, Valiukonyte M, et al. Transcriptomic and genetic analysis suggests a role for mitochondrial dysregulation in schizophrenia [Internet]. medRxiv; 2025 [cited 2025 May 9]. p. 2025.03.14.25323827. Available from: https://www.medrxiv.org/content/10.1101/2025.03.14.25323827v1

38. Tkachev A, Stekolshchikova E, Vanyushkina A, Zhang H, Morozova A, Zozulya S, et al. Lipid Alteration Signature in the Blood Plasma of Individuals With Schizophrenia, Depression, and Bipolar Disorder. JAMA Psychiatry. 2023 Mar 1;80(3):250–9.

39. Xu W, Zhang D, Ma Y, Gaspar RC, Kahn M, Nasiri A, et al. Ceramide synthesis inhibitors prevent lipid-induced insulin resistance through the DAG-PKCε-insulin receptorT1150 phosphorylation pathway. Cell Rep. 2024 Oct 22;43(10):114746.

40. O’Gallagher K, Teo JTH, Shah AM, Gaughran F. Interaction Between Race, Ethnicity, Severe Mental Illness, and Cardiovascular Disease. J Am Heart Assoc. 2022 Jun 21;11(12):e025621.

