## Supplement for "MRI-derived body composition in bipolar and psychotic disorders"

**Supplementary information**

| **Variable** | **CTR (n=251)** | | **BD&PD (n=63)** | | **BD**  **(n=36)** | | **PD**  **(n=27)** | | **CTR vs. BD&PD** | | **CTR vs.**  **BD** | | **CTR vs.**  **PD** | |
| --- | --- | --- | --- | --- | --- | --- | --- | --- | --- | --- | --- | --- | --- | --- |
|  | **Mean** | **SD** | **Mean** | **SD** | **Mean** | **SD** | **Mean** | **SD** | **T** | **P** | **T** | **P** | **T** | **p** |
| Glucose | 5.2 | 0.6 | 5.2 | 0.5 | 5.1 | 0.5 | 5.2 | 0.4 | -0.0 | 0.983 | 0.3 | 0.771 | -0.4 | 0.675 |
| HbA1c | 32.0 | 3.2 | 32.3 | 3.7 | 31.8 | 3.8 | 32.8 | 3.5 | -0.3 | 0.728 | 0.3 | 0.787 | -1.0 | 0.329 |
| CRP | 1.7 | 2.8 | 3.1 | 6.4 | 3.4 | 8.1 | 2.6 | 3.2 | -1.7 | 0.101 | -1.3 | 0.217 | -1.5 | 0.151 |
| HDL-C | 1.6 | 0.5 | 1.5 | 0.5 | 1.6 | 0.6 | 1.4 | 0.5 | 1.3 | 0.192 | 0.3 | 0.796 | 2.0 | 0.053 |
| LDL-C | 3.2 | 0.8 | 2.9 | 0.8 | 3.1 | 0.8 | 2.7 | 0.7 | 2.1 | 0.035 | 0.6 | 0.526 | 3.0 | 0.005 |
| Total-C | 5.0 | 1.0 | 4.8 | 0.9 | 5.0 | 0.9 | 4.5 | 0.7 | 1.6 | 0.120 | -0.0 | 0.985 | 3.2 | 0.003 |
| TG | 1.1 | 0.7 | 1.1 | 0.5 | 1.0 | 0.5 | 1.2 | 0.6 | -0.0 | 0.993 | 0.5 | 0.617 | -0.6 | 0.566 |

**Table S1: Sample characteristics.** Blood serum values and tests for differences (two-sided t-tests) between controls (CTR) and all patients (BD&PD) and bipolar disorder (BD) and psychotic disorder (PD). *Abbreviations:* HbA1c=glycated hemoglobin, HDL-C=high-density lipoprotein cholesterol, LDL-C=low-density lipoprotein cholesterol, Total-C=total cholesterol, TG=triglycerides. *Missing data points:* CTR: CRP (n=99), Glucose (n=99), HbA1c (n=200), HDL-C (n=99), LDL-C (n=99), TG (n=150); BD: CRP (n=2), glucose (n=2), HbA1c (n=2), HDL-C (n=2), LDL-C (n=2), Total-C (n=2), TG (n=2); PD: CRP (n=1), glucose (n=1), HbA1c (n=1), HDL-C (n=1), LDL-C (n=1), Total-C (n=1), TG (n=1).

| **Variable** | **BD (n=36)** | | **PD (n=27)** | | **BD vs. PD** | |
| --- | --- | --- | --- | --- | --- | --- |
|  | **Mean/N** | **SD/%** | **Mean/N** | **SD/%** | **T/χ2** | **p** |
| Age | 36.6 | 10.9 | 31.7 | 8.9 | 2.0 | 0.054 |
| Female | 23 | 63.9 | 9 | 33.3 | 4.6 | 0.032 |
| BMI | 26.2 | 3.8 | 26.0 | 4.3 | 0.2 | 0.828 |
| Underweight | 0 | 0 | 0 | 0 |  |  |
| Normal weight | 14 | 38.9 | 12 | 44.4 | 0.0 | 0.827 |
| Overweight | 16 | 44.4 | 9 | 33.3 | 0.4 | 0.543 |
| Obese | 5 | 13.9 | 5 | 18.5 | 0.0 | 0.868 |
| Glucose | 5.1 | 0.5 | 5.2 | 0.4 | -0.6 | 0.568 |
| HbA1c | 31.8 | 3.8 | 32.8 | 3.5 | -1.1 | 0.286 |
| CRP | 3.4 | 8.1 | 2.6 | 3.2 | 0.5 | 0.602 |
| HDL-C | 1.6 | 0.6 | 1.4 | 0.5 | 1.3 | 0.201 |
| LDL-C | 3.1 | 0.8 | 2.7 | 0.7 | 1.9 | 0.061 |
| Total-C | 5.0 | 0.9 | 4.5 | 0.7 | 2.4 | 0.020 |
| TG | 1.0 | 0.5 | 1.2 | 0.6 | -1.0 | 0.341 |
| ASAT | 7.1 | 3.0 | 6.3 | 3.5 | 0.9 | 0.354 |
| VAT | 2.7 | 1.8 | 2.8 | 1.9 | -0.2 | 0.877 |
| Liver fat | 3.0 | 2.0 | 2.4 | 1.0 | 1.8 | 0.081 |
| MFI | 6.8 | 1.8 | 6.2 | 1.8 | 1.5 | 0.140 |
| Muscle volume | 2.7 | 0.6 | 3.1 | 0.7 | -2.0 | 0.050 |

**Table S2: Patient sample characteristics.** Characteristics of the patient sample and tests for differences between bipolar disorder (BD) and psychotic disorder (PD). We used t-tests for continuous and chi-squared tests for categorical measures. *Abbreviations*: BMI=body mass index, HbA1c=glycated hemoglobin, HDL-C=high-density lipoprotein cholesterol, LDL-C=low-density lipoprotein cholesterol, Total-C=total cholesterol, TG=triglycerides, ASAT=abdominal subcutaneous adipose tissue, MFI=muscle fat infiltration. *Missing data points:* BD: BMI (n=1), CRP (n=2), glucose (n=2), HbA1c (n=2), HDL-C (n=2), LDL-C (n=2), Total-C (n=2), TG (n=2), MFI (n=1), muscle volume (n=1); PD: BMI (n=1), CRP (n=1), glucose (n=1), HbA1c (n=1), HDL-C (n=1), LDL-C (n=1), Total-C (n=1), TG (n=1), LF (n=3).

| **Outcome** | **β** | **SE** | **T** | **r** | **95% CI** | **p** |
| --- | --- | --- | --- | --- | --- | --- |
| **Antipsychotic dose** | | | | | | |
| BMI | 0.04 | 0.03 | 1.42 | 0.19 | -0.07, 0.42 | 0.203 |
| ASAT | 0.16 | 0.09 | 1.74 | 0.23 | -0.02, 0.45 | 0.138 |
| VAT | 0.16 | 0.08 | 2.02 | 0.26 | 0.01, 0.48 | 0.099 |
| LF | -0.17 | 9.68 | -0.02 | -0.00 | -0.25, 0.25 | 0.986 |
| MFI | 0.01 | 0.00 | 1.78 | 0.23 | -0.02, 0.46 | 0.138 |
| MV | 0.09 | 0.09 | 1.05 | 0.14 | -0.11, 0.38 | 0.335 |
| **Antipsychotic dose, verified** | | | | | | |
| BMI | 0.03 | 0.03 | 1.06 | 0.16 | -0.13, 0.41 | 0.335 |
| ASAT | 0.15 | 0.10 | 1.52 | 0.22 | -0.06, 0.47 | 0.185 |
| VAT | 0.16 | 0.09 | 1.89 | 0.27 | -0.00, 0.51 | 0.122 |
| LF | 2.05 | 10.30 | 0.20 | 0.03 | -0.25, 0.30 | 0.883 |
| MFI | 0.01 | 0.00 | 1.74 | 0.25 | -0.03, 0.49 | 0.138 |
| MV | 0.11 | 0.09 | 1.23 | 0.18 | -0.10, 0.43 | 0.265 |

**Table S3: Associations between antipsychotic dose and body composition measures.** Results from multiple linear regression analyses on the associations of antipsychotic dose with body mass index (BMI), abdominal subcutaneous adipose tissue (ASAT), visceral adipose tissue (VAT), liver fat (LF), muscle fat infiltration (MFI), and muscle volume (MV) adjusted for age and sex. The first analyses were performed in all patients, the second without those whose antipsychotic use could not be verified by serum concentration measures. P-values are adjusted with the Benjamini-Hochberg method. Statistically significant adjusted p-values are in bold. *Abbreviations*: β=regression coefficient, SE=standard error, T=t-statistic, r=partial correlation coefficient, CI=confidence interval, P=p-value.

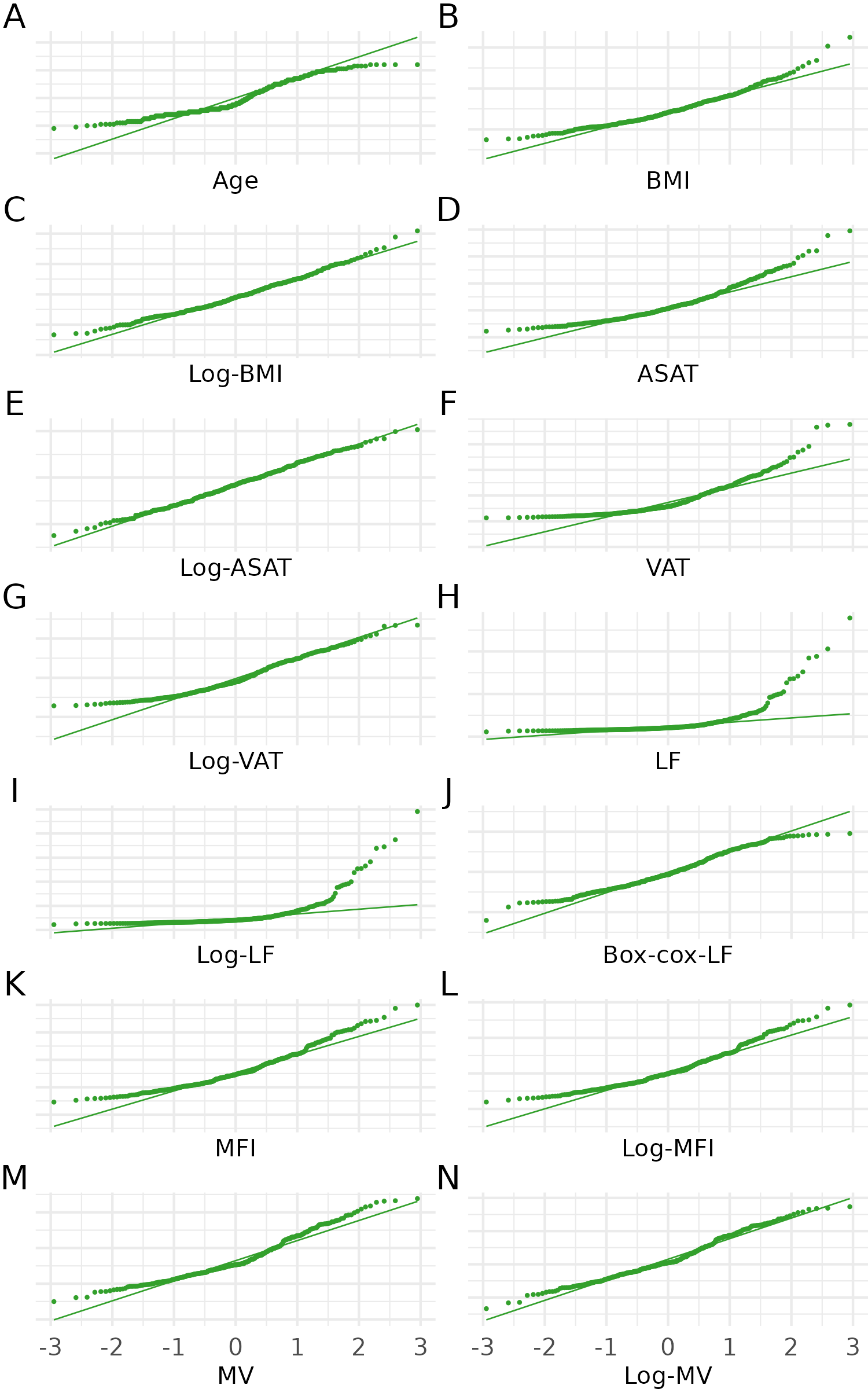

**Figure S1: Quantile-quantile plots of age, anthropometric and body composition measures.** Quantile-quantile plots of age, body mass index (BMI), abdominal subcutaneous adipose tissue (ASAT), visceral adipose tissue (VAT), liver fat (LF), muscle fat infiltration (MFI), and muscle volume (MV), when relevant transformed.

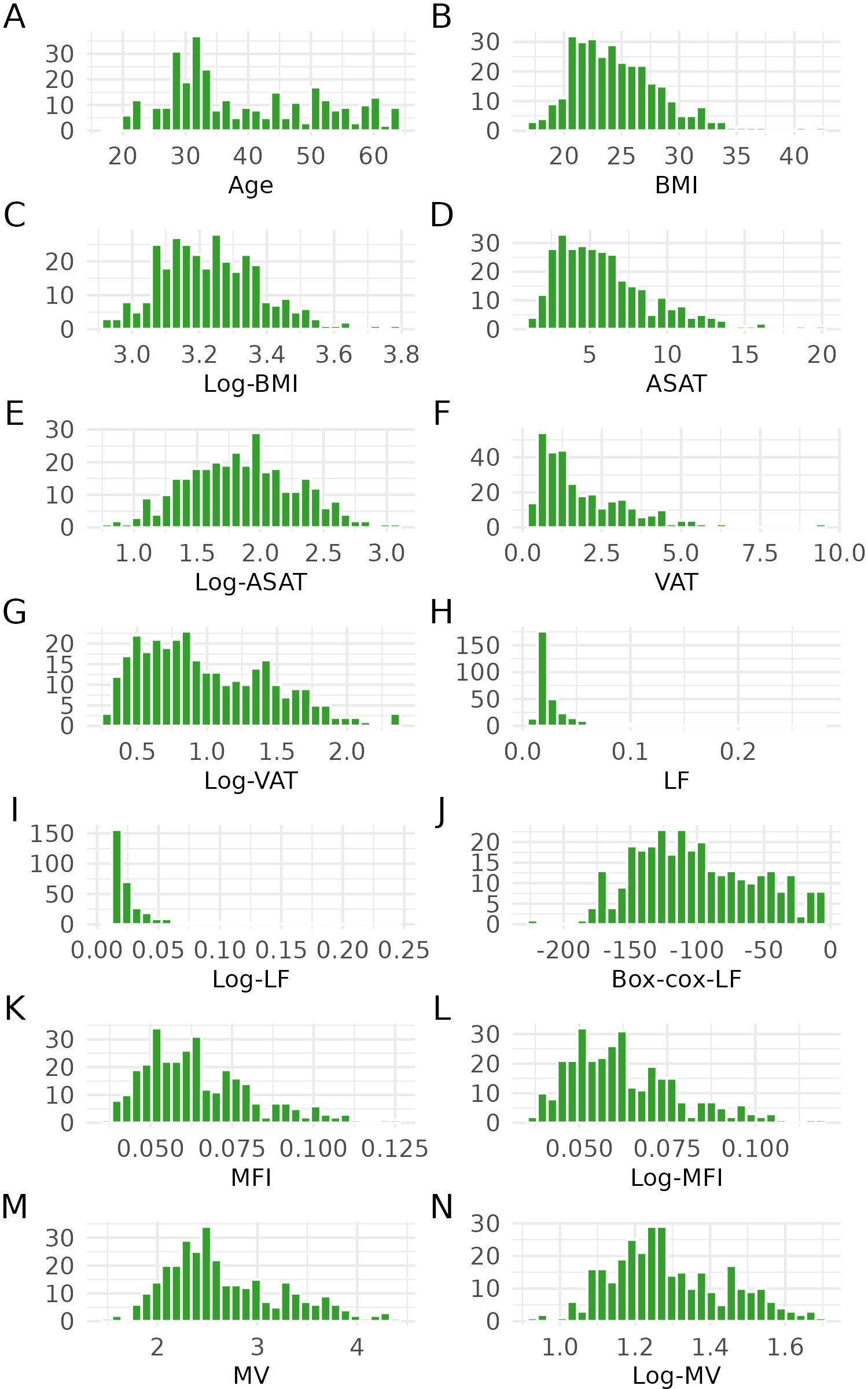

**Figure S2: Histograms of age, anthropometric and body composition measures.** Histograms of age, body mass index (BMI), abdominal subcutaneous adipose tissue (ASAT), visceral adipose tissue (VAT), liver fat (LF), muscle fat infiltration (MFI), and muscle volume (MV), when relevant transformed.
